# Predicting hospital demand during the COVID-19 outbreak in Bogotá, Colombia

**DOI:** 10.1101/2020.04.14.20065466

**Authors:** Claudia Rivera-Rodriguez, Piedad Urdinola

**Author notes:** Correspondence: Claudia Rivera-Rodriguez.

## Abstract

Colombia, like many developing nations, does not have a strong health system able to respond to a pandemic of the magnitude of Covid-19. There is an increasing need to create a model that allows particular clinics and hospitals to estimate the number of patients that require Intensive Care Units-ICU care (critical), and the number of patients that require hospital care (severe), but not ICU care, in order to manage their limited resources.

This paper presents a prediction of the total number of ICU and regular beds that will be needed during the pandemic COVID-19 for Bogotá-Colombia. We use a SEIR model that includes three different compartments of infection: those who can stay at home, those in regular hospital beds and those in need of ICU treatment. The model allows for a time varying transmission rate which we use to incorporate the measures introduced by the government over the period of one semester. The model predicts that by mid October 2020, the city will need 4 524 prevalent ICUs needed and 16 738 regular hospital beds needed. By the third week of July 2020, the number of patients that need ICUs will overpass the capacity set at 1 200 beds for ICU hospital beds in the city. The model predicts that the death toll by the same date will reach 1 752 people and the number of cases will be 30 216 inhabitants by then. We provide a Shiny app available in https://claudia-rivera-rodriguez.shinyapps.io/shinyappcovidclinic/. The original values in the app reproduce the results of this paper, but the parameters and starting values can be changed according to the users needs. COVID-19 has posed too many challenges to health systems around the globe, this model is an useful tool for cities, hospitals and clinics in Colombia that need to prepare for the excess demand of services that a pandemic like this one generates. Unfortunately, the model predicts that by the third week of July the projected capacity of the system in Bogotá will not be enough. We expect the lockdown rules strength in the future days, so the death toll is not as bad as predicted by this model.

## 1 BACKGROUND

The novel coronavirus disease 2019 (COVID-19) epidemic has spread from China to almost all countries in the world by April 1st 2020. The first official case is reported in Colombia on March 6, 2020 from an imported case and evolved to local cases of transmission. In order to reduce the impact of the COVID-19 outbreak in Bogotá, the largest city in Colombia, a local lockdown was introduced on March 15, 2020, followed by a national lockdown on March 19, 2020. Colombia, like many developing nations, does not have a strong health system able to respond to a pandemic of the magnitude of the present one. Neither in terms of infrastructure, medical personnel nor in terms of logistic preparedness or technical capacities to arrange all medical needed resources. The latter is the main motivation to create a model that allows particular clinics and hospitals to estimate the number of beds and the respirators needed to attend during the peak days. Specifically, we are interested in estimating the number of patients that require Intensive Care Units-ICU care (critical), and the number of patients that require hospital care (severe), but not ICU care.

As of April 04, 2020, Colombia had only carried out 460 test per million people (https://infogram.com/, https://ourworldindata.org/covid-testing), whereas other countries such as Germany and South Korea have done over 1000 test per million people. Additionally, on March 26 one of the two available machines used to run the detection tests broke, leading to a reduction on its production and causing delays in the total number of cases detected. Unfortunately, for developing countries like Colombia it has been an enormous effort to expand facilities and production of biotechnology inputs to run the necessary number of tests required to detect all active cases of the virus, the highest number up to date is 17 000 tests on June 19/2020. Thus, one of the biggest concerns is that the data may not inform well how many hospital beds (and ICU beds) will be needed during the outbreak peak. In fact, one of the main caveats for this study is that the official data is very likely to be under estimated as only patients with at least one symptom or that have had contact with another detected case are being tested (INS). Moreover, we are assuming an overall probability of requiring ICU treatment, while sex, age and co-morbidity (diabetes, hypertension, acute respiratory diseases and depressed immune system) will highly determine differential probabilities, that we are not taken into account.

We implemented a SEIR model (Susceptible - Exposed - Infectious - Recovered) to forecast the number of cases in Bogotá, the largest city in Colombia and the one with the most numbers of cases to date, using the public official COVID-19 information from Health Secretariat-Saludata and available at (). The model includes three different compartments of infection: Infected that require ICU care, Infected that require hospital care, but not ICU care and Infected that only require Home care. The model accounts for the effect of control strategies introduced by the government by changing the transmission rate over time. We developed a Shiny app that displays the results from the model publicly available at (https://claudia-rivera-rodriguez.shinyapps.io/shinyappcovidclinic/). The initial parameters are the ones used in this work, users can change the parameters according to their needs. The Shiny app can works as a forecast tool for individual clinics by specifying the market share (percentage) of the population corresponding to the clinic. During the outbreak some clinics should be ready to see an increase in their market share because they may have more resources, such as ICU beds and the model allows for each clinic to adjust it. The model can be used for specific cities or towns, the user only needs to change the population size, and some parameters of interest.

The results from our assumptions show that Bogotá city will need over 2000 beds, the Mayor’s target number of ICUs, by the end of May 2020, as this goal was not reached it was modified to a lower number of 1 200 ICU beds by mid August. By the last week of March there were 300 beds available and by June 20 there are 766 available ICU beds for COVID-19 patients. This number could be well below Bogotá’s needs based on our projections.

## 2 METHODS

SIR methods (Susceptible-Infected-Recovered) became widespread in the prediction of communicable diseases since its creation in the early 20th century (Anderson, 1991). Several authors have provided forecasting models using this method as presented in Murray (2020), but SIR models heavily rely on initial strong assumptions. SEIR models are a variation that relaxes some of those assumptions, including closed populations and account for communicable diseases that transmit in transitions, starting from the entire population (Susceptible) that incubate the disease for a period of time (Exposed) making the person infected but not infectious (I) and finally become Recovered (R) (Martcheva, 2015). Each transition holds a rate based on what is observed from a population, that is a susceptible person gets infected at a transmission rate once in contact with an infected individual, and become exposed. Once exposed transition to infected happens at a rate that captures the inverse of the mean latent period of the disease. The final transition is recovery with permanent immunity. We chose this model to estimate bed demands per institution in Colombia separating regular and ICU (Intensive Care Units) beds, which allows a distinction for each type that reflects differential transition rates, preparedness and logistics for health providers. Similar works have been used to forecast similar needs in Europe and United States of America (Li et al., 2020; Massonnaud et al., 2020; Zhang et al., 2020a) and more recently they are also part of the discussion in developing nations, SIR models are also used to forecast the virus progression in Colombia (Manrique et al., 2020).

## 3 MODEL

We fitted a deterministic SEIR model over 6 months, for practical purposes it is important to bear in mind that policies change along this time scenario and therefore the models must be updated. The population is divided into Susceptible (S), Exposed (E), Infected (I), recovered (R), and D (death). Those Infected (I) are subdivided into three compartments: *I*_U_, *I*_NoU_, *I*_H_ which, respectively, denote infected individuals that require ICU care, infected individuals that require hospital care, but not ICU, and infected individuals that only require home care.

One implication of our model is that it does not consider events such as births, and it only considers deaths due to COVID-19. Note that we assume that patients transit from E to ICU care directly, therefore we assume that the average time from (E) to (*I*_U_) is larger than the average time from (E) to (*I*_NoU_) and subsequently this is larger than the average time from (E) to (*I*_H_). These transitions and considerations are summarized in figure 1. We also assume that only patients in ICU die, while other infected patients recover. The total population of Bogotá is 7.4 million, but we assume an initial population size of 8 million to account for its metropolitan area because people commute to work and study daily from those surrounding towns to Bogoá Capital District.

**Figure 1.**
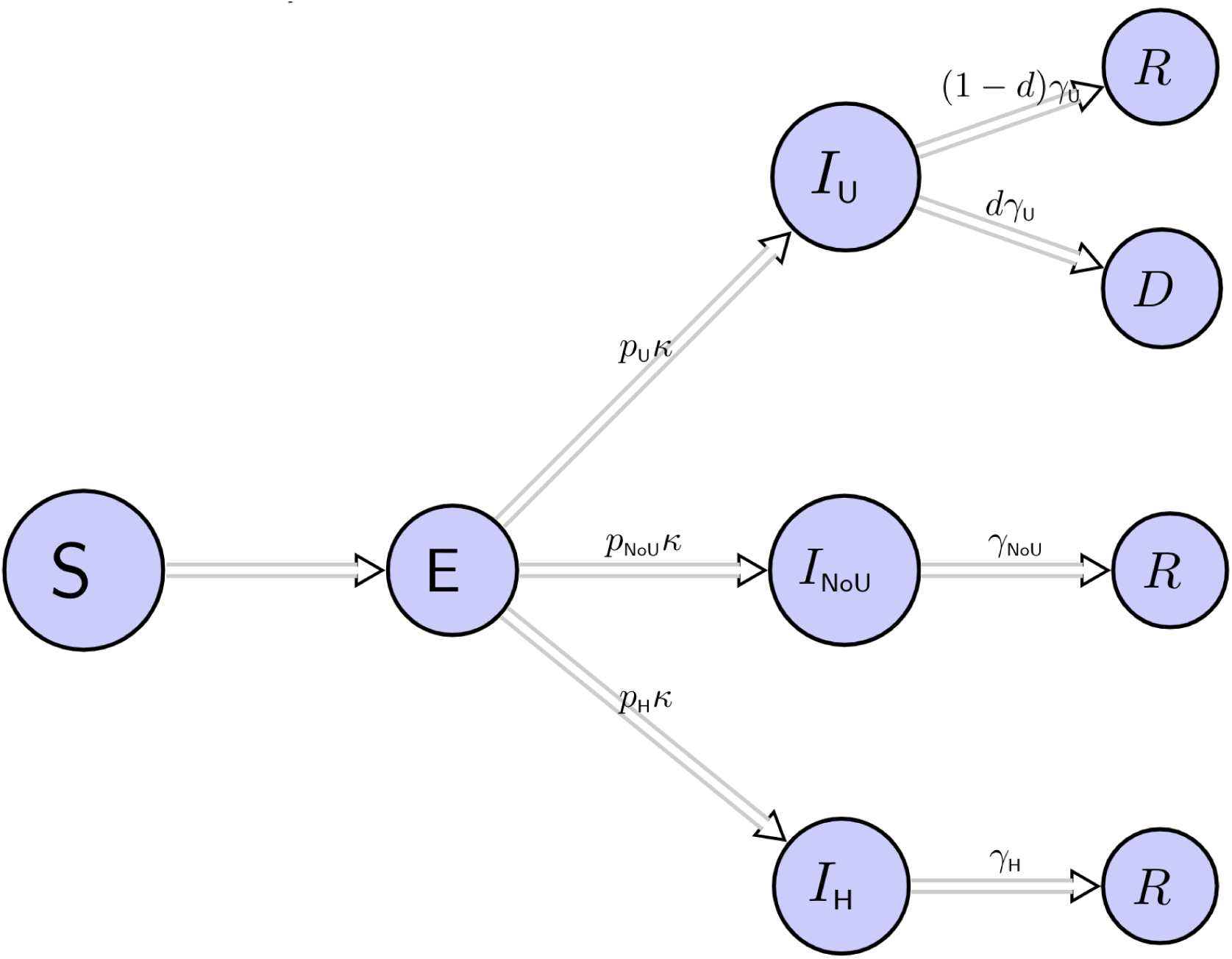
We divided the population into susceptible (*S*), exposed (*E*), infected ICU (*I*_U_), infected in hospital but not ICU (*I*_NoU_), infected that require only home care (*I*_H_), recovered (*R*) and dead subjects (*D*) individuals. Infected subjects are *I*_U_, *I*_NoU_ or *I*_H_ with probabilities *p*_U_, *p*_NoU_ and *p*_H_ respectively. The term 1*/κ* is the mean incubation period and 1*/γ*_U_, 1*/γ*_NoU_, 1*/γ*_H_ are the daily probabilities that the respective patients recover. *d* is the probability of death for ICU patients.

We describe the epidemic transitions through the model

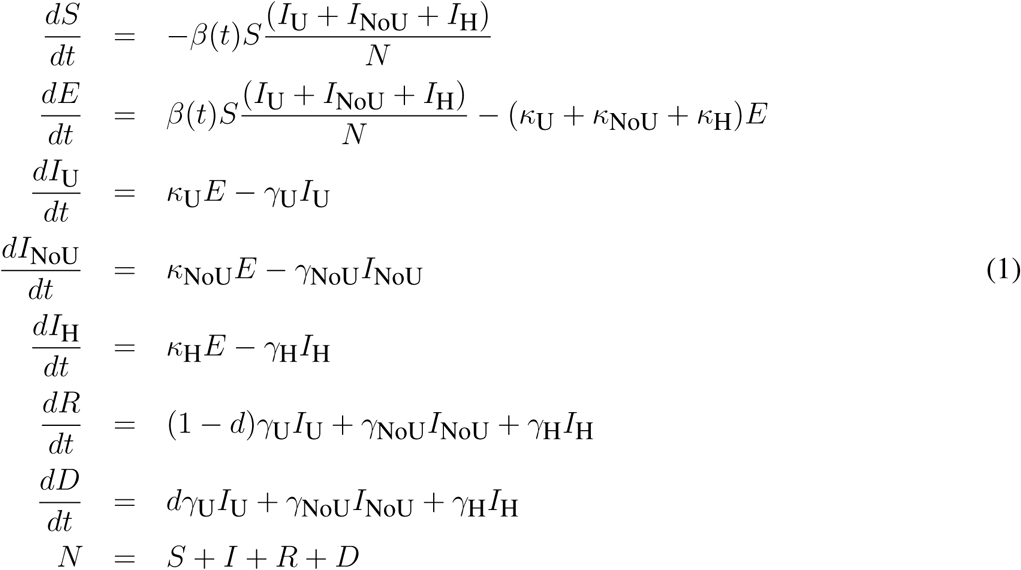

where *κ*NoU = *p*NoU*κ, κ*U = *p*U*κ* and *κ*H = *p*H*κ*. To model the impact of interventions introduced by the government, we allow the transmission rate to be a step-wise function *β*(*t*), with 3 steps at *t*_0_, *t*_1_ and *t*_2_. The time *t*_0_(2020-05-24) corresponds to the case when we start predicting, *t*_1_(2020-06-16) is when new measures are introduced and *t*_2_ (2020-06-30) is the date when measures are revised and implemented. We estimate *β*(*t*_0_) from the basic reproduction number such that *R*_0_ = 1.1 (Bog, 2020). For *t* > *t*_0_, we choose *β*(*t*) such that *R*(*t*) ≈ 1.3, for *t*_1_ ≤ *t* < *t*_2_ and *R*(*t*) ≈ 1.2, for *t* ≥ *t*_2_ (Bog, 2020).

The terms *p*U, *p*NoU and *p*H denote the probabilities that case requires ICU care, hospital non-ICU care and only home care, respectively. Note that *p*U + *p*NoU + *p*H = 1. To estimate these probabilities, we use information from the Colombian National Health Institute, finding *p*U = 0.0168, *p*NoU = 0.14 and *p*H = 0.843. The parameter *κ* is the daily probability of an exposed individual becoming infected, and *γ*U, *γ*NoU, *γ*H are the daily probabilities of the corresponding infected individuals becoming recovered or dead. The probability *d* denotes the probability that an infected ICU individual dies. Table 1 displays the parameters of the models, their interpretation and sources The starting values for the model are based on the numbers from Bogotá, Colombia reported by May 24. There where 7166 cases, 1318 recovered and 212 deaths by then in the city.

**Table 1.** Parameters and definition for model (1)

| Symbol | Definition | Value | Source |
| --- | --- | --- | --- |
| $\beta(t)$ | Transmission rate | Stepwise function | (Liu et al., 2020a; Kucharski et al., 2020) |
| $\kappa$ | Daily probability of an exposed individual becoming infected: $\kappa = 1/\alpha$, with $\alpha$ being the mean incubation period | 1/5.2 | (Zhang et al., 2020b) |
| $p_U$ | Probability of patient being ICU | 0.0168 | (Bog, 2020) |
| $p_{NoU}$ | Probability of patient being in Hospital, but not ICU | 0.14 | (Bog, 2020) |
| $p_H$ | Probability of patient being mild/at home | 0.843 | (Bog, 2020) |
| $\gamma_U$ | Daily probability that an infected individual in ICU recovers, when the mean infection period is $b_U$, $\gamma_U = 1/b_U$ | 1/6 | (Liu et al., 2020b; Zhang et al., 2020b; Prem et al., 2020) |
| $\gamma_{NoU}$ | Daily probability that an infected individual in Hospital, but no ICU recovers, when the mean infection period is $b_{NoU}$, $\gamma_{NoU} = 1/b_{NoU}$ | 1/5 | (Zhang et al., 2020b; Prem et al., 2020; Lin et al., 2020; Liu et al., 2020b) |
| $\gamma_H$ | Daily probability that an infected individual in Hospital, but no ICU recovers, when the mean infection period is $b_H$, $\gamma_H = 1/b_H$ | 1/5 | (Zhang et al., 2020b; Prem et al., 2020; Lin et al., 2020; Liu et al., 2020b) |
| $d$ | Probability of dying given that patient is in ICU | 0.50 | (Wu and McGoogan, 2020) |

## 4 RESULTS

Figure 2 shows the results for each category the model predicts, ICU and regular hospital beds. Even all the positive measures assumed in the model, we predict that the peak of the epidemic can happen around 2020-11-14. During the epidemic peak, the model predicts that 1 362 ICUs will be needed for coronavirus patients, and 9 470 non-ICU hospital beds. We predict that the maximum number of prevalent cases will be 67866. With the parameters in the models, the total number of deaths could reach 13 268 in 6 months time.

**Figure 2.**
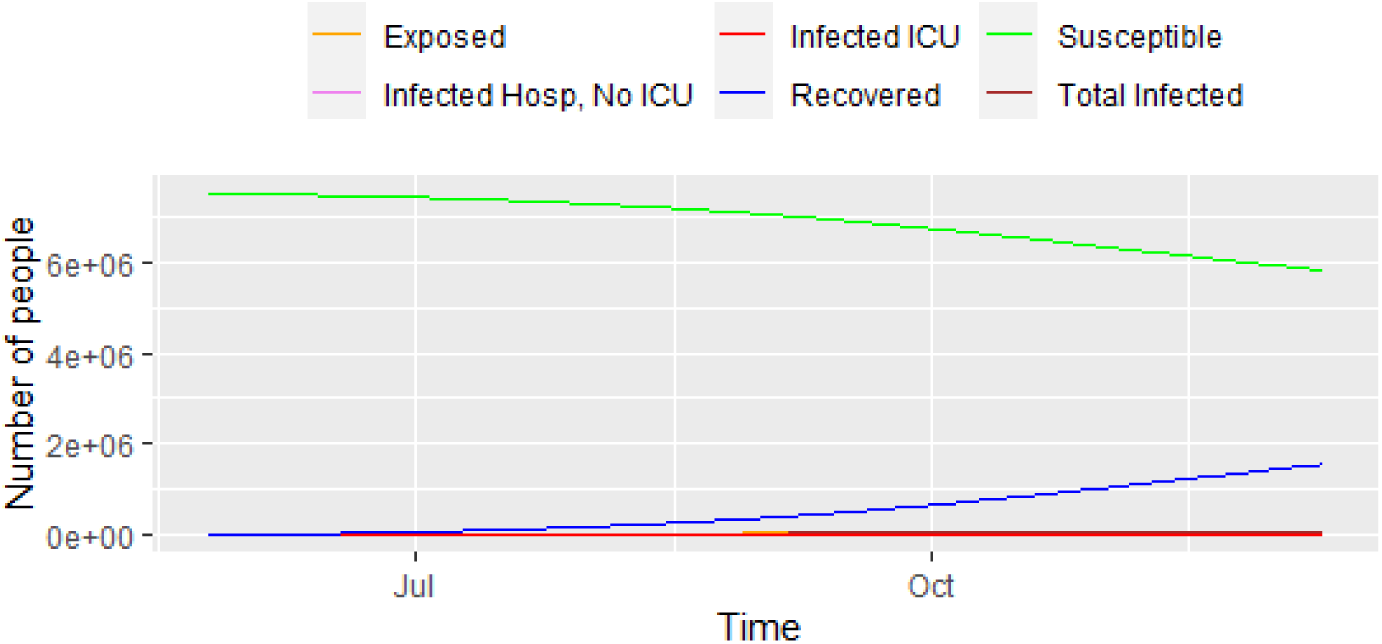
Progress of the epidemic using model (1)

Figure 3 displays a closer picture of those infected and the total number of deaths. We can see that the total number of infected that will need hospital care (ICU and non-ICU) are concernedly high. Additionally, figure 4 shows those infected that will need hospital care, compared to the current number of ICU beds in the city. It shows that the number of ICUs needed will be 1362, i.e. the city has to increase its capacity in order to provide care to everyone that needs it. The local authorities in Bogotá are planning to have a total of 1 200 ICU beds in the city, but the current number is still lower than that. The number 1 200 is overrun by mid September/2020, with a death toll of 4 850 people by then. Unfortunately, the trend keeps on increasing for the following months, which reflects the lack of preparedness for a catastrophe like the current one in Bogotá and likely in other similar developing nations.

**Figure 3.**
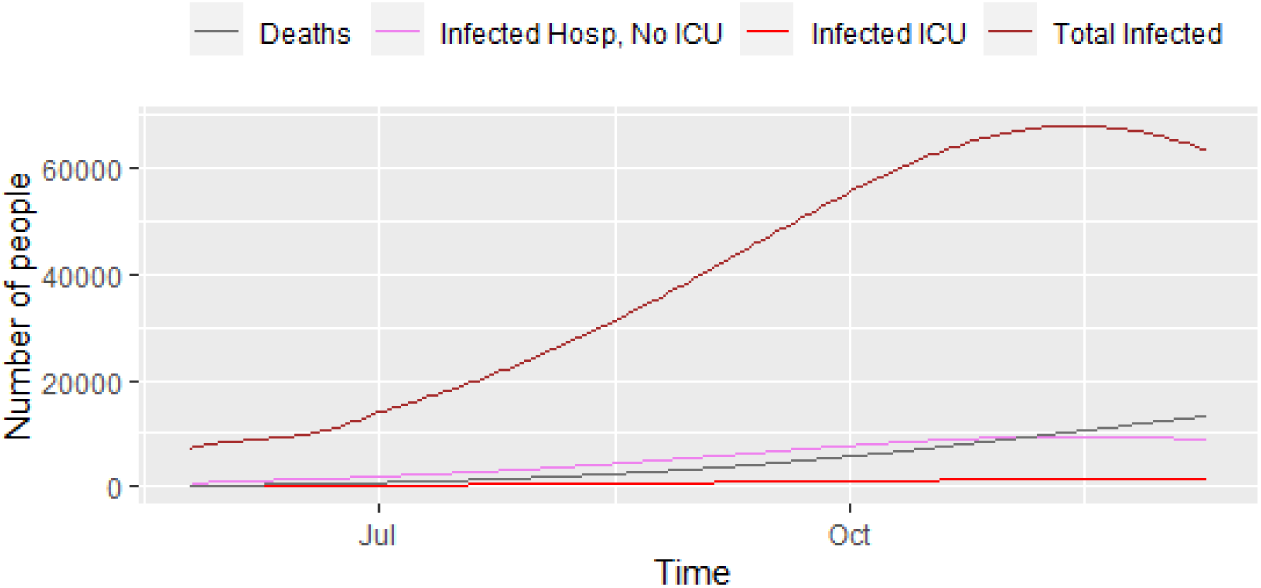
Progress of the epidemic - infected and deaths, results from model (1)

**Figure 4.**
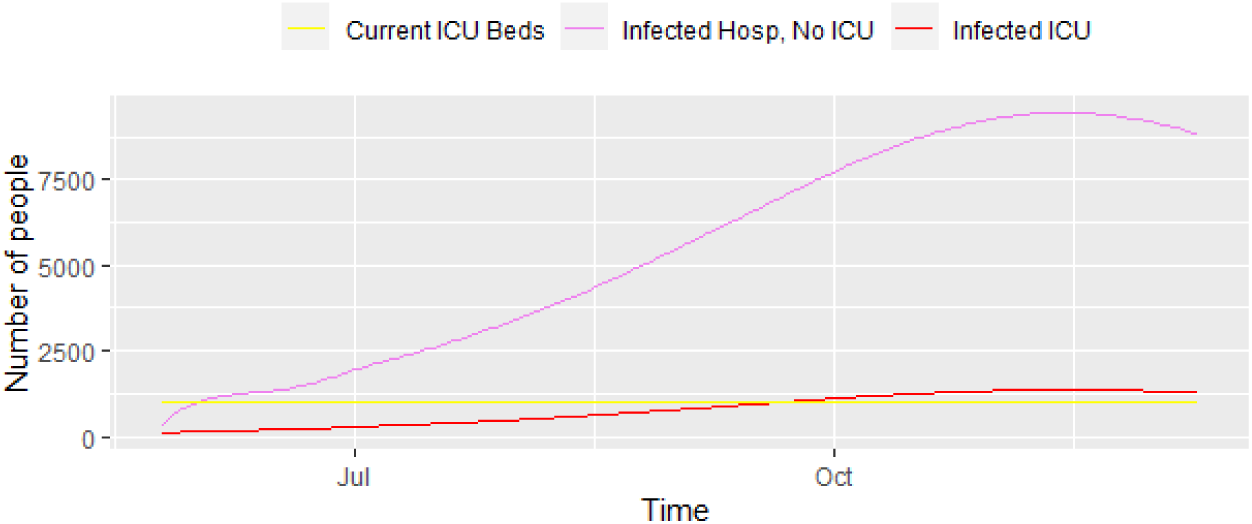
Progress of the epidemic - Infected that require ICU and infected that require regular hospital beds, results from model (1)

When we double the number of days of ICU patients to recover in the model, i.e 1*/γ*U = 14 days, rather than 7, the number of ICU beds almost double in size. Moreover, if the probability of being an ICU patient *p*U is reduce, the number of beds is reduced, but even an small increase in this probability will cause a large increase in the number of ICU beds needed during the peak of the epidemic.

## 5 DISCUSSION

This document presents a prediction of the total number of ICU and regular beds the would be needed during the pandemic COVID-19 for Bogotá-Colombia. We use a SEIR model that differentiates between three types of infected patients: those who can stay at home, those in regular hospital beds and those in need of ICU treatment. With a mean incubation period of 5.2 days and mean infection period of 4 days (for patients at home), 5 days (for patients in regular hospital beds) and 7 days (for patients in ICU). The parameters assumed in the model are for a positive scenario, where the effective reproduction number during the lock-down is assumed to be 1.5, and 2 after the lock-down, when other measures are introduced. We assume that 2.6% of patients require ICU treatment, 13.4% require regular hospital beds, and the rest only require home care. The model allows for a time varying transmission rate which we use to incorporate the measures introduced by the government over the period of 1 year. The model predicts that by mid July, 2020, the city will reach the peak of the epidemic with a total of 22 541 active ICU prevalent cases and 85 015 regular hospital beds needed.

By the end of May 2020, the number of patients that need hospitalization will overpass the current planned capacity set at 1200 beds for ICU beds in the city by mid August, and the death toll will reach a total of 1752. Unpreparedness of the health system will only increase COVID-19 related and unrelated mortality, as already observed in Italy, USA and other countries. Measures like lockdown have been used in most countries to diffuse the demand for health services due to COVID-19 over time, however it may be insufficient if there are not enough resources to amp the health services in developing nations, such is the case of Colombia, where the need of additional resources are a priority at this point.

Other than the intrinsic limitations of SEIR models, this prediction model does not account for age and sex distribution of the population but we plan to introduce such differences in a future version of the model with an additional mixing including the contact matrices, as the recently national population census in Colombia is available. Also, we have fitted a a model with two interventions: a lockdown and mitigation measures, but this can be modified later in time. Neither we accounted for regional differences that in a tropical context relate to weather and climate, because there is no evidence, to date, that the novel coronavirus could or not spread in an homogeneous pattern under certain weather conditions.

Finally, we provide a Shiny app available in https://claudia-rivera-rodriguez.shinyapps.io/s The original values in the app reproduce the results of this paper, but the parameters and starting values can be changed according to the users needs.

## 6 CONCLUSIONS

COVID-19 has posed too many challenges to health systems around the globe, this model is an useful tool for cities, hospitals and clinics in Colombia that need to prepare for the excess demand of services that a pandemic like this one generates. Unfortunately, the model predicts that by July capacity of the system in Bogotá will not be enough. We expect the lock-down rules strength in the future days, so the death toll is not as bad as predicted by this model.

## Data Availability

This is a simulation study, code is available through a shiny app.

https://claudia-rivera-rodriguez.shinyapps.io/shinyappcovidclinic/

## ABBREVIATIONS

SEIR: Susceptible-Exposed-Infected-Recovered
ICU: Intensive Care Unit
COVID-19: Coronavirus disease 2019

## DECLARATIONS

### Ethics approval and consent to participate

Not applicable

### Consent for publication

Not applicable

### Availability of data and materials

The materials(code) used and/or analysed during the current study are available from the corresponding author on reasonable request. Additionally, a shiny app is available online at https://claudia-rivera-rodriguez.shinyapps.io/shinyappcovidclinic/

### Authors’ contributions

CRR contributed to the analysis and coding of the model and the shiny app. PU contributed to model interpretation and article writing. All authors have read and approved the manuscript.

## Acknowledgements

We thank the Colombia research group COVID Modelling CG for their feedback.

## 1 REPRODUCTION NUMBER AND *β*(*T*)

We use the next generation matrix approach to find the basic reproduction number (van den Driessche, 2017; Diekmann et al., 2009). We estimate *β*(*t*_0_) from the basic reproduction number. To find *R*_0_, following Martcheva (2015), we let **x** = [*E, I*U, *I*_NoU_, *I*_*H*_] and **y** = [*S, R, D*]. Disease free equilibrium is **x**_0_ = [0, 0, 0, 0] and **y**_0_ = [*N*, 0, 0]. Then,

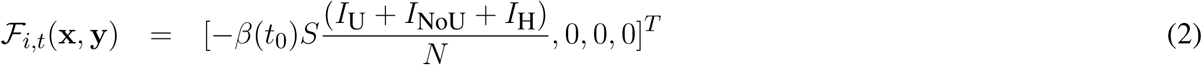

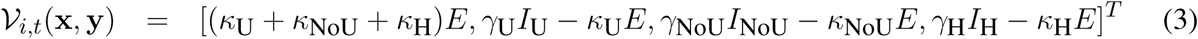

and

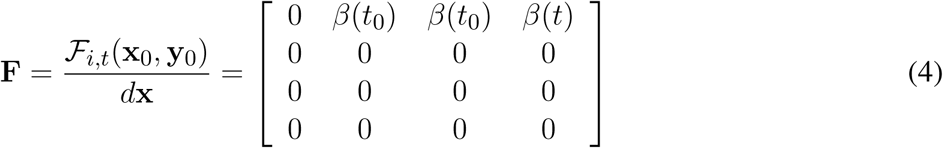

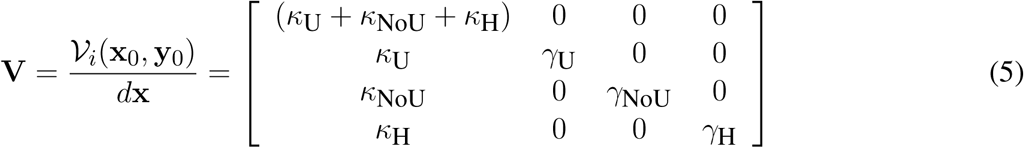

The inverse of **V** is given by

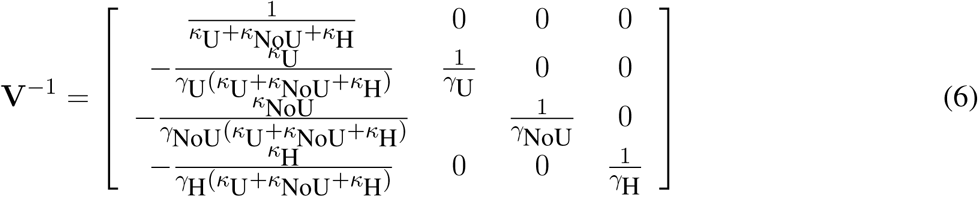

thus

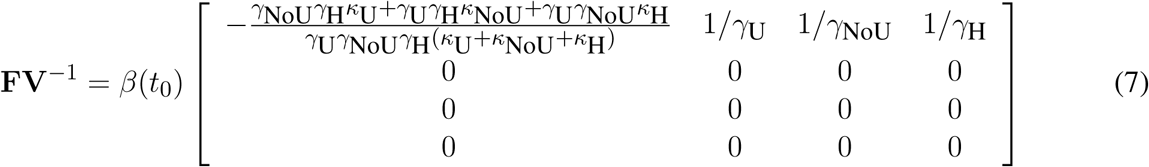

The spectral radio of **FV**^−1^ is

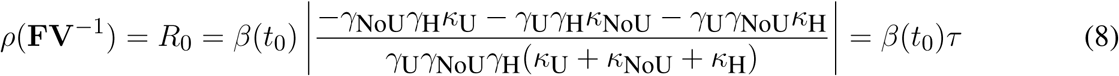

Using this, we find that

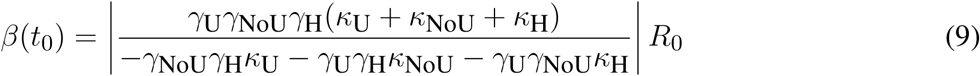

When *R*_0_ = 1.1, *β*(*t*_0_) = 0.22. Our rational for choosing *β*(*t*_1_) and *β*(*t*_2_) is as follows.

Using the initial *β*_0_, we calculate *S*(*t*_1_)*/N*, and we assume that *R*(*t*) = 1.3 for *t*_1_ ≤ *t* ≤ *t*_2_, and *R*(*t*) = 1.1 for *t* > *t*_2_.

To find *β*(*t*), we assume that *R*(*t*) ≈ *β*(*t*) ∗ *τ S*(*t*)*/N*. So, we have that *β*(*t*) ≈ *R*(*t*) ∗ *N/*(*τ* ∗ *S*(*t*)) So, we have *β*(*t*) = 0.26 for *t*_1_ ≤ *t* ≤ *t*_2_, and *β*(*t*) = 0.24 for *t* > *t*_2_.

